# Proof-of-Concept Validation of Noninvasive Detection of Cortical Spreading Depolarization with High Resolution Direct Current-Electroencephalography with Future Device Recommendations

**DOI:** 10.1101/2024.11.12.24311133

**Authors:** Benjamin R. Brown, Samuel J. Hund, Kirk A. Easley, Eric L. Singer, C. William Shuttleworth, Andrew P. Carlson, Stephen C. Jones

## Abstract

**Background/Objective:** Cortical spreading depolarization (SD) is increasingly recognized as a major contributor to secondary brain injury. Noninvasive SD monitoring would enable the institution of SD-based therapeutics. Our primary objective is to establish proof-of-concept validation that scalp DC-potentials can provide noninvasive SD detection by comparing scalp direct-current (DC)-shifts from a high-density electrode array to SDs detected by gold-standard electrocorticography (ECoG). Our secondary objective is to assess usability and artifact tolerance.

**Methods:** An 83x58 mm thermoplastic elastomer array with 29 6-mm diameter Ag/AgCl 1-cm spaced electrodes, the CerebroPatch™ Proof-of-Concept Prototype, was adhesively placed on the forehead with an intervening electrode gel interface to record DC-electroencephalography in normal volunteers and severe acute brain injury patients in the neuro-intensive care unit some with and some without invasive ECoG electrodes. The scalp and ECoG voltages were collected by a Moberg® Advanced ICU Amplifier. Artifacts were visually identified and usability issues were recorded. SD was scored on ECoG based on DC-shifts with associated high-frequency suppression and propagation. A six-parameter Gaussian plus quadratic baseline model was used to estimate ECoG and scalp electrode time-courses and scalp-voltage heat-map movies. The similarity of the noninvasive scalp and invasive ECoG DC-shift time-courses was compared via the Gaussian fit parameters and confirmed if the Coefficient-of-Determination was >0.80.

**Results:** Usability and artifact issues obscured most scalp Prototype device data of the 140 ECoG-coded SDs during 11 days in one sub-arachnoid hemorrhage patient. Twenty-six of these DC-shifts were in readable, artifact-free portions of scalp recordings and 24 of these had a >0.80 Coefficient-of-Determination (0.98[0.02], median[IQR]) between invasive ECoG and noninvasive Prototype device DC-shifts. Reconstructed heat-map movies of the scalp DC-potentials showed a 5-cm extent, -460 µV peak region that persisted for ∼70 sec. These data suggest that these scalp DC-shifts (peak -457±69 µV [mean±StD], full-width-half maximum 70.9±5.92 sec, area 18.7±2.76 cm^2^) depicted in the heat-map movies represent noninvasively detected SDs.

**Conclusions:** These results using 26 SDs as the observational units suggest that noninvasive SD detection is possible using scalp DC-potential signals with a high spatial resolution EEG array. Although the high artifact burden data and low usability records were limiting, negative results, they serve as an important entrepreneurial recipe that provides suggestions for a future, re-designed device that would reduce artifacts and improve usability for DC-EEG SD detection needed to enable multi-modal monitoring for secondary brain injury.

## 1. Introduction

Cortical spreading depolarizations (SDs) are slowly traveling regions of depolarized neurons and glial cells that travel as waves in the cerebral gray matter in which evoked and spontaneous electroencephalography (EEG) are suppressed [1–4]. This en masse depolarized region causes a negative swing in field potential of 5 to 30 mV [5, 6] that is reduced by ∼90% at the scalp surface [6, 7]. SDs spread at a rate of 1-9 mm/min across the cortical surface [1, 2, 8], thus taking tens of minutes to travel across the human brain surface. In humans, SDs have been found to follow severe acute brain injuries (sABIs) [3, 4], including malignant hemispheric stroke, severe traumatic brain injury, and sub-arachnoid hemorrhage (SAH). SDs are also associated with migraine auras [4, 9, 10], concussion [7, 11, 12], and high-grade malignant glioma [13–15].

During the 2-week extended period in the neuro-intensive care unit (ICU) following sABI, SDs have been strongly implicated as a factor in secondary brain injury [16]. This pathogenic process of secondary brain injury [17, 18] results in increased brain damage, increased mortality and morbidity, and more extensive rehabilitation. Many clinical studies by the CoOperative Study on Brain Injury Depolarizations (COSBID) group have shown that SDs: 1) increase the area of necrosis [8, 19–23]; 2) are associated with unfavorable outcomes in severe traumatic brain injury [22, 24]; and 3) can be used to predict outcome and initiative rescue measures in aneurysmal SAH (aSAH) [25]. Recently, the duration of SD-induced EEG suppression has been associated with infarct progression in malignant hemispheric stroke patients [26]. These results suggest an urgent need for SD-based therapeutics to mitigate SD’s damaging effects. Answering this need are several positive pilot studies using ketamine [27] and nimodipine [28] and a comprehensive review of other agents [29]. These positive results [27, 28] need confirmation in large clinical trials using noninvasive SD monitoring based on the estimated twenty times the number of closed-skull compared to craniotomy sABI patients that would be available using invasive SD monitoring. This invasive method involves the direct placement of electrocorticography (ECoG) electrode strips onto the cortical surface [30, 31] requiring either a craniotomy [21] or an enlarged burr hole [21, 32] and is considered the gold standard with its three COSBID-defined identification criteria that include a propagating DC-shift and its associated EEG suppression [33, 34]. Alternatively, depth electrodes can be used to detect SD either placed horizontally and subdurally in an extraventricular drainage burr hole [35, 36] or inserted intracortically via a multimodal monitoring bolt placed at the bedside [37, 38].

This need for noninvasive SD detection, expressed repeatedly [4, 24, 33] but starting with SD’s first demonstration in humans by Strong et al. [31], has inspired several efforts directed at scalp SD detection: one attempt demonstrated scalp direct current (DC)-shifts and EEG suppression but not SD propagation so was partially successful [6]. Other efforts used decompressive craniotomy patients with a portion of their skulls removed that provided overly favorable conditions but showed some promise in such patients [39–41]. Another noninvasive attempt based on Hartings et al. [39] but without ECoG was unsuccessful [42, 43]. Recently a case study reported retrospectively visually identified decreases in EEG delta band (0-4 Hz) power that were time-associated with depth electrode ECoG suppressions and DC shifts of a SD [44], suggesting another possible method of noninvasive SD detection. Another study demonstrated scalp EEG detection of SDs associated with epileptiform field potentials [45]. All three SD-identification aspects of the gold standard COSBID criteria [33, 34] were successfully recorded, but this study did not provide ground truth confirmation [45]. Of course, artifacts occur in all of these methods so must be addressed accordingly [46].

Simulation studies of both the primary DC-shift [7] and the secondary EEG suppression [47] attributes of SD have been used to better define effective methods for noninvasive detection. One simulation effort used silent cortical dipoles imbedded in a realistic brain model to simulate 0.5-40 Hz EEG suppression to explore the well-known variation in suppression duration and width as a determinant of detectability [47]. Hund et al. [7] used finite element analysis based on Poisson’s equation to perform numerical simulations that estimated the scalp DC-potentials from a concussive brain surface SD modeled as an expanding ring. Hund et al. [7] then used these estimates of brain-surface DC-potential to explore the optimal electrode configuration for reliable scalp SD detection and showed that closely-spaced electrodes are a necessary component for faithful noninvasive detection using DC-EEG.

We have implemented Hund et al.’s [7] design recommendations for noninvasive SD detection in the forehead-placed CerebroPatch™ Proof-Of-Concept Prototype (see Figure 1). Here, we present proof-of-concept validation data showing the close correspondence between ECoG-identified SDs and Prototype-device-detected slow potential changes characterized by DC-shifts that occur over a period of about 3 minutes as well as artifact and usability data. Our hypothesis is that DC-potentials recorded by closely-spaced scalp electrodes can be associated with ECoG-detected SDs and that these scalp DC-potentials provide noninvasive SD detection but that artifact and usability issues must be addressed in an improved electrode array design.

**Figure 1:**
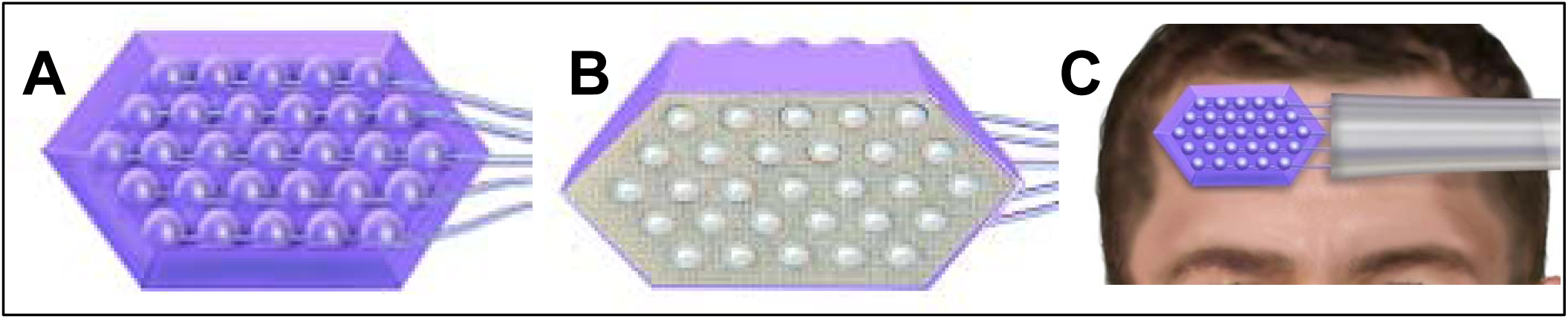
CerebroPatch™ Proof-of-Concept Prototype: A, Top view showing the domes covering the 29 1-cm spaced Ag/AgCl electrodes with their attached conductors; B, Angled bottom view showing double-sided adhesive and gelled electrode pits; C, Target position of Prototype device on forehead.

## 2. Methods

### 2.1 CerebroPatch™ Proof-of-Concept Prototype description

The CerebroPatch™ Proof-of-Concept Prototype electrode array (see Figure 1) is a single-layer 83x58 mm thermoplastic elastomer scaffold (Dynaflex™ G2706-1000-00, Avient Corporation, Avon Lake, OH) encasing a 1-cm spaced hexagonal array of 29 6-mm diameter 1.3-mm thick sintered Ag/AgCl electrodes (BMD-6, Biomed Products Inc., Fair Oaks, CA) with a 0.5 mm lip such that the active diameter is reduced to 5 mm. The array was fixed on the forehead with a double-sided adhesive membrane (3M 1522 Medical Double Coated Tape, 3M Corporation, Saint Paul, MN) cut to conform to the array outline and 5-mm electrode openings. Electrode gel (Ten20® Conductive Paste, Weaver and Company, Aurora, CO) filled the 1-mm space between the Ag/AgCl electrodes and the skin after the skin was prepped with Lemon Prep™ (Mavidon Corp., Flat Rock, NC) and cleaned with sterile water.

### 2.2 Human study issues

The CerebroPatch™ Proof-of-Concept Prototype was classed as a Non-Significant Risk device during the protocol’s evaluation and ethical approval by the Institutional Review Board of the University of New Mexico Human Research Protections Office (UNM HRPO 18-051). All ethical guidelines have been adhered to, the proper ethical approvals were obtained from the institutional review board, and self or surrogate informed consent was obtained from the participants or a legally authorized representative.

### 2.3 Protocol Details

This proof-of-concept study consisted of three cohorts with five subjects each: Cohort 1 consisted of normal subjects; Cohort 2 included neuro-ICU patients with ischemic stroke, intracranial hemorrhage, severe traumatic brain injury, or SAH who were implanted with a 1x6 ECoG strip electrode (AU1X6P Auragen 1x6 Cortical Strip Platinum, Integra LifeSciences Corporation, Plainsboro, NJ) placed at the time of craniotomy for their sABIs; and Cohort 3 were similar patients to Cohort 2 but with intact skulls. In Cohort 1 patients the CerebroPatch™ Proof-of-Concept Prototype device was placed on the forehead. In Cohort 2 patients the Prototype device was placed on the forehead over the 6-electrode ECoG strip. No direct confirmation of their co-registration was possible as the Prototype device was removed for imaging procedures and every 24 hours to record a skin irritation score. It was then re-gelled and reapplied. The skin irritation score consisted of the sum of scores for erythema and edema reactions as in ISO 10993-10 [48; see Table 1]. In Cohort 3 patients the Prototype device was placed on the forehead over the presumed edge of the lesion. A mastoid Ag/AgCl reference electrode was used in all cases. Cohort 1 participants were monitored for 2 hours in an effort to assess drift due to the transepithelial potential [49, 50] that could then be used to correct patient data. In Cohorts 2 and 3 monitoring was continued for up to 14 days to allow for the maximal number of SDs.

**Table 1.**
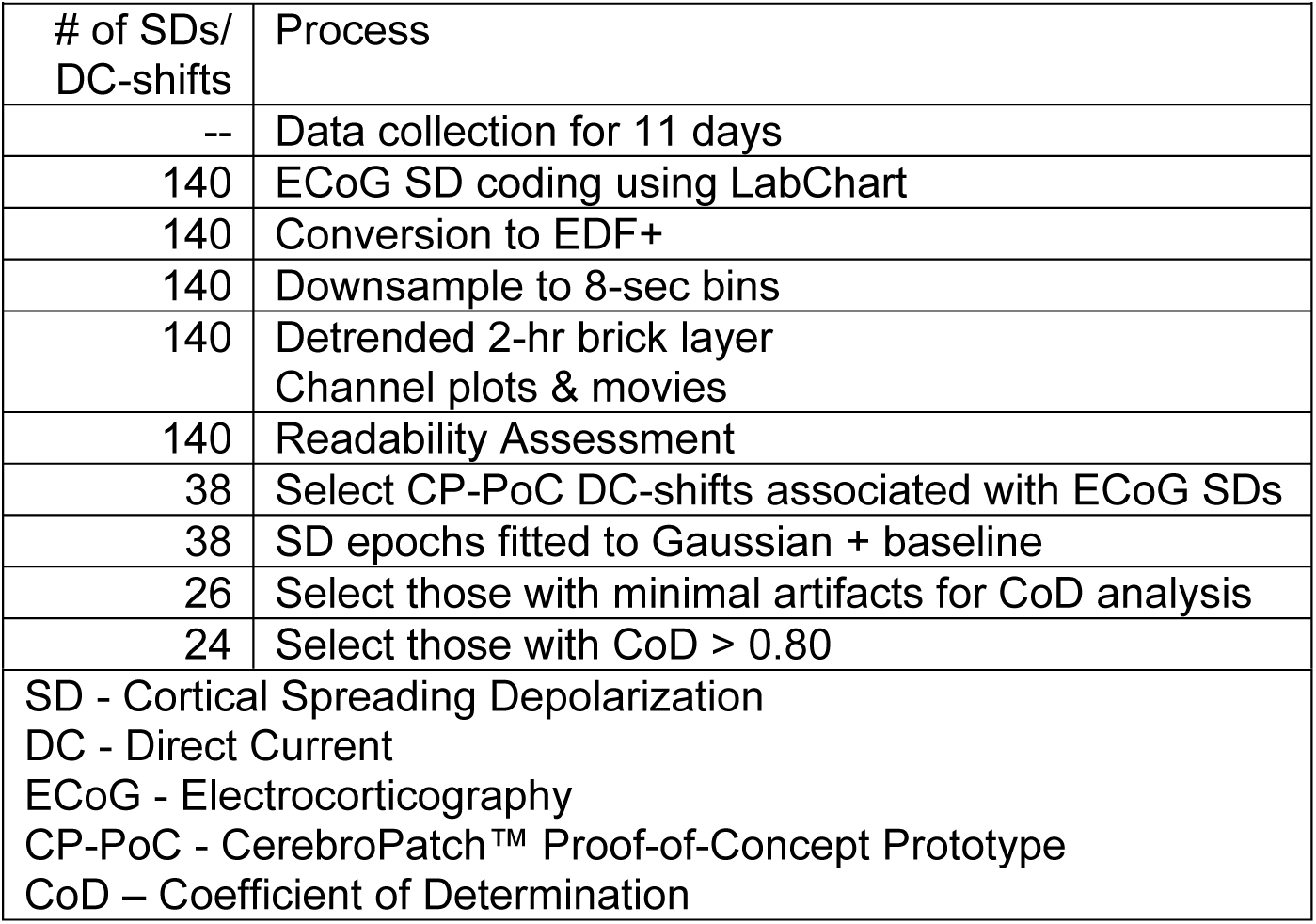
Signal Processing Workflow.

### 2.4 Data collection, ECoG SD coding, and data analysis

Table 1 presents the signal processing workflow as described in detail below.

Voltages from the CerebroPatch™ Proof-of-Concept Prototype and the ECoG strip were collected by a DC-coupled Moberg® CNS Advanced ICU Amplifier (Moberg Research Inc., Micromed Group, Ambler, PA) at 256 Hz. ECoG recordings were coded for SDs and isoelectric SDs (ISDs) using standard COSBID criteria [33, 34] by the study site principal investigator, APC, blinded to Prototype device data, using LabChart software (ADInstruments, Inc., Colorado Springs CO, USA). Only times with valid ECoG recording were scored. Briefly, SDs were identified as a propagating characteristic DC-potential shift coupled with suppression of clinical-frequency (0.5-50 Hz) ECoG data. Those SDs that occurred repetitively in the same location [33, 46] following a previous definite SD were “scored even if partially obscured by artifacts or other recording problems” [51]. This criteria of repetition has been shown to improve ECoG scoring of SDs in high-artifact recordings [46]. The duration of ECoG suppression was also recorded. If a characteristic DC-shift occurred in a period when the EEG signal was absent indicating isoelectric tissue, then it was coded as an ISD.

For further unblinded analysis, the brain-surface ECoG and scalp Prototype device signals were converted to EDF+ format [52] and processed using lab-designed Python scripts. The 256 Hz data was pre-processed in all 29 channels of the Prototype device and 6 channels of the ECoG strip by determining the mean voltage of the DC signal within non-overlapping 8 second segments, reducing the signal sample rate to 0.125 Hz, consistent with oversampling by a factor of ∼20 based on the estimated SD frequency of 0.006 Hz.

Detrended time-course channel plots of ECoG and CerebroPatch™ Proof-of-Concept Prototype v oltages were generated after the data were segmented into overlapping 2-hr “brick-layer” epochs (to minimize edge effects from the detrending procedure). The detrending procedure consisted of fitting a 2-hr epoch to a linear function, and then subtracting this function from the data, to provide a linear baseline over the 2-hr data block. These data sets were then used to generate heat-map movies of the Prototype device data for visualization of the scalp electric field. This visualization involved interpolating the voltage between the sensors via Gaussian-based radial basis functions [53, 54] using a square grid with 0.33 mm spacing. A radial Gaussian profile of width equal to the sensor spacing (1 cm) was used without smoothing. Missing data from dead channels was filled by this spatially-aware interpolation process.

Readable portions of CerebroPatch™ Proof-of-Concept Prototype channel plots with minimal artifacts were visually assessed by SCJ. Recording periods were classed as unreadable based on the presence of any of the following:

1. Excessive and large amplitude artifacts with frequencies that obscured SDs;
2. DC-deflections that occurred in all Prototype device channels at the same time;
3. A high auto-scale range (above 1000 µV) indicative of artifacts; or
4. A low scale range (below 50 µV) indicative of a “no recording” period.

The scale range was chosen by excluding the most extreme 1% of the values to display just the middle 99% of the data.

### 2.5 Criteria of identifying scalp DC-shifts that are associated with ECoG-coded SDs

DC-shifts from the CerebroPatch™ Proof-of-Concept Prototype heat-map movies and both of the overlapping 2-hr brick-layer channel plots were classed as acceptable for comparison with the ECoG DC-shifts from coded SDs using the following criteria 1-3, and 4 if appropriate:

1. A DC-shift of varying amplitude occurred in more than one channel, but not all channels, as observed both in the movies and channel plots. For instance, Figure 2B shows the varying amplitude of channels B1 through B6 and the lack of signal in channels A1, C1, C2, and D3.
2. The magnitude of the DC-shifts were between -230 to -1200 µV. This is consistent with the range of 7% [7] and 12% (unpublished data) estimated and 3% experimental [7] scalp/brain-surface voltage ratios applied to the 1^st^ and 3^rd^ quartiles of ECoG amplitudes of -7.2 mV to -10.1 mV [7] as derived retrospectively from Drenckhahn et al.’s [6] data as reported by Hund et al. [7].
3. The spatial extent estimated from the DC-shift movies was ∼4 cm to be consistent with the estimated x12 spatial spread of the ECoG DC-shifts [7].
4. A DC-shift occurs repetitively in the same location. This repetition criteria is based on the COSBID criteria for identifying SDs from Dreier et al. [33; p.22; last paragraph: ‘similar pattern of DC shift/SPC as recorded at a different time’]. These criteria have recently been shown to improve ECoG scoring of SDs in high-artifact recordings [46].

**Figure 2:**
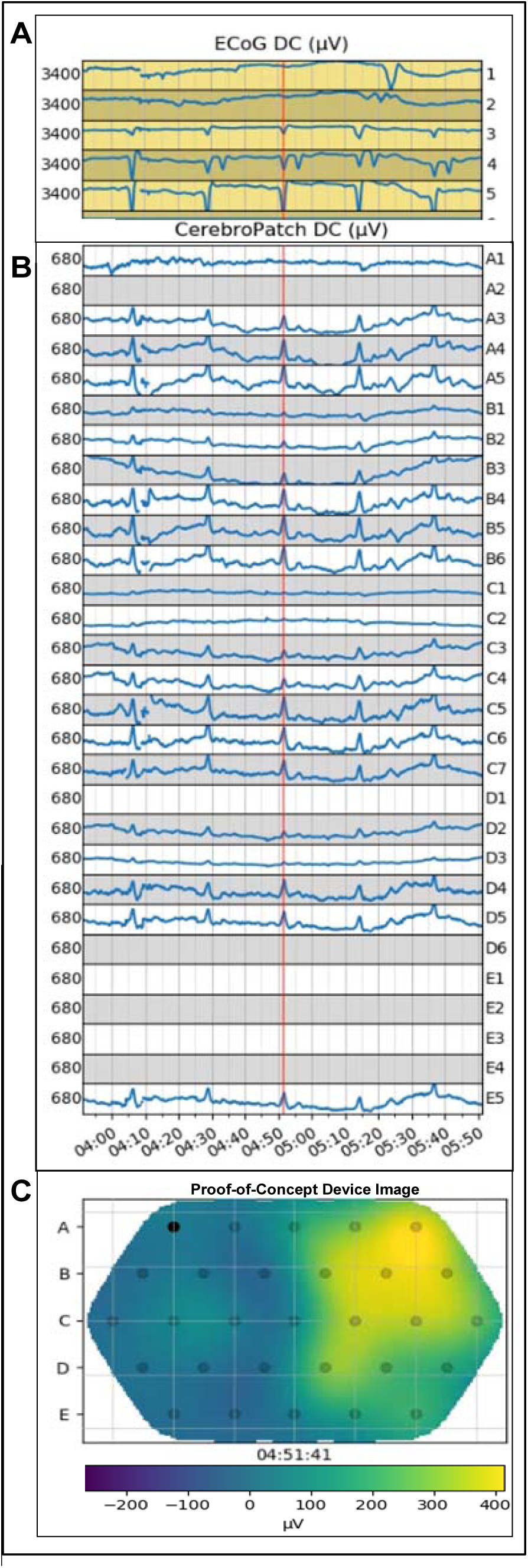
The ECoG strip channel plots (Panel A, 3400 µV full-scale, negative down) show the DC-shifts from coded-SDs in channels 3, 4, and 5 (and 2 and 1 for the 4^th^ SD) with the electrode labels along right edge. CerebroPatch™ Proof-of-Concept Prototype (Panel B, 680 µV full scale, negative up) channel plots (dead channels A2, D1, D6, E1-4 data removed) show 5 time-matched DC-shifts from the forehead-placed Prototype device showing their temporal synchrony. The red vertical time marker is at the peak of the 3rd SD at 4:51. These SDs are part of the cluster of 25 SDs that have an inter-SD interval of 22 min in which SDs occurred in the same channels in a repetitive pattern. Panel C shows the Prototype device image at the 4:51 peak of the 3^rd^ SD showing the extent of the SD at the scalp. The electrodes in Panel C are represented as small black circles labeled in five rows with 5 electrodes in rows A and E, 6 in rows B and D, and 7 in row C with a filled-circle depicting electrode A1. In the bottom panel the pseudo-color scale ranges from 250 to -410 µV (positive-negative reversed as negative is “up”).

This process resulted in the selection of Prototype device channel data that could be compared with time-matched ECoG DC-shifts.

### 2.6 Scalp and ECoG DC-shift curve fitting, comparison, visualization, and spatial extent

The scalp and ECoG data was processed via a curve fitting procedure designed to provide equivalent flat baselines and create curve-fitting parameters to define and quantitatively compare their time-courses. Given that the two signals from the ECoG strip and CerebroPatch™ Proof-of-Concept Prototype device were collected with different sensors and with different background interferences, this processing enables them to be quantitatively compared on an “equal” basis, as if there were recorded under exactly the same conditions. This processing involved subtracting a fitted baseline, fitting each data set to a Gaussian, and scaling the Prototype device data with the ratio of the maximum voltage extents as detailed below. This procedure allowed the quantitative comparison, not only of the fitted parameters as shown in Table 2, but also the comparison of time-courses via the square of the correlation coefficient, the Coefficient-of-Determination (CoD).

**Table 2.** Comparison of CerebroPatch™ Proof-of-Concept Prototype with ECoG DC-shift curve fit parameters.

|  |  |  |  |  |  |  |
| --- | --- | --- | --- | --- | --- | --- |
|  |  | Signal origin |  |  |  |  |
| Parameter | Units | CerebroPatch™<br>Proof-of-Concept<br>Prototype device | ECoG | Ratio or<br>Difference | Fraction of<br>duration | n |
| Peak Voltage | ($\mu\text{V}$) | $-457 \pm 69$ | $-3771 \pm 171$ | $0.121 \pm 0.016$ | - | 24 |
| Duration<br>(FWHM)* | (sec) | $70.9 \pm 5.92$ | $63.2 \pm 3.65$ | $1.12 \pm 0.076$ | - | 24 |
| Frequency | (Hz) | $0.0063 \pm 0.0005$ | $0.0070 \pm 0.0004$ | - | - | 24 |
| Peak Time<br>Difference** | (sec) | - | - | 2.89 [2.32] | 4.3% | 23 |
| Area*** | ($\text{cm}^2$) | $18.7 \pm 2.71$ | | | | 24 |
| Diameter**** | (cm) | $4.86 \pm 0.358$ | - | - | - | 24 |
| Coefficient-of-Determination<br>($>0.80$)***** | (-) | - | - | $0.98 [0.02]$ | - | 24 |
| Values are mean ± StD or median [interquartile range, IQR] | | | | | | |
| Thirteen of the 39 epochs were excluded from analysis due to lack of readability and 2 out of the remaining 26 for Coefficient-of-Determination criteria |  |  |  |  |  |  |
| * $p < 0.001$, paired t-test | | | | | | |
| ** With only scalp delayed (excluding one preceding) |  |  |  |  |  |  |
| *** Area within the 50% isocontour of maximum voltage extent |  |  |  |  |  |  |
| **** Diameter of a circle with an area within the 50% isocontour |  |  |  |  |  |  |
| ***** Based on 8-sec data averages |  |  |  |  |  |  |

To implement this quantitative comparison of the Prototype device readable DC-shifts and ECoG DC-shifts coded as SDs, an epoch width that captured the entire DC-shift and included sufficient regions of stable baseline was chosen. The average DC-voltages in the 8 second segments within each of these epochs of the 29 Prototype device and 6 ECoG channels were fitted using least squares to estimate the following six parameters: three parameters for a Gaussian function model characterized by amplitude, width (or ’sigma’), and the maximum signal time and three parameters for a quadratic baseline consisting of a DC offset, a DC linear drift, and a second-order DC drift. The ECoG and Prototype device channels with the maximum amplitude (largest deviations from zero) were used to compare their respective peak DC-shifts. The ratio of the maximum amplitudes was used to scale the 8 second binned Prototype device signals to that of the ECoG signal to compute the CoD for the comparison of their structural similarity. A CoD value of >0.80 was deemed as suggesting acceptable similarity. The Gaussian parameters of signal amplitude, time of the peak signal, the full-width-half-maximum (FWHM), as 2.355 x sigma, and frequency as 1/FWHM were used to compare the Prototype device and ECoG DC-shifts.

The spatial extent of the scalp voltage field detected by the Prototype device from the brain-surface SDs was estimated as the area of the contiguous region above the 50% isocontour of the DC-voltage amplitude at the peak voltage time. For comparison with our previous [7] and unpublished simulation results, the diameter of a circle with the equivalent area was calculated.

### 2.7 Statistics

Statistical tests were performed using SAS/STAT version 15.1 (SAS institute Inc, Cary, NC). Parametric or nonparametric statistical tests were used appropriately depending on the Shapiro-Wilk test for normality. Outliers were not excluded as SDs vary widely depending on their surrounding metabolic and circulatory milieu. Different measures of central tendency are reported depending on the Shapiro-Wilk test: the mean ± standard deviation (StD) or the median [interquartile range, IQR]. A 2-sided p value of less than 0.05 was accepted as statistically significant.

## 3. Results

### 3.1 Skin irritation scores, study interruptions and usability

The only instance of the skin irritation scores being other than zero was a score of 2 (erythema) recorded after 6 days for one subject. The CerebroPatch™ Proof-of-Concept Prototype was removed and data collection halted. In one subject, skin depressions (“bumps”) at the electrode sites were noted during the re-gelling procedure but subsided over an hour. For another subject, the Prototype device was removed and data collection halted after 7 days due to family complaints. There were multiple adhesion issues reported in 7 out of 10 patients with many comments in the data sheets describing difficulties due to the device not sticking to the scalp after its removal and re-application for daily skin irritation assessment and for imaging procedures.

### 3.2 Joint ECoG and CerebroPatch™ Proof-of-Concept Prototype analysis

None of the CerebroPatch™ Proof-of-Concept Prototype data from Cohorts 1 and 3 was available due to excessive artifacts, which obscured any attempts in Cohort 1 at evaluating the drift due to the transepithelial potential. Out of the five subjects in Cohort 2, two had no identifiable ECoG SDs and two had excessive artifacts in the Prototype device signals. Data analysis focused on one 66-to-70-year-old female aSAH patient, whose aneurysm was clipped, with 101 SDs and 39 ISDs identified from the LabChart analysis of the ECoG data over a period of 11 days at which time the patient was discharged to in-patient rehabilitation. In this 11 day period, there were 0.85 days of readable Prototype device data with 1.84 days of no recording and 8.31 days of unreadable Prototype device signals. All 140 of these LabChart identified SDs and ISDs were visualized using the Python script depiction of the ECoG voltage time series. From this analysis, these ECoG-coded SDs and ISDs exhibited multi-ECoG electrode DC-shifts primarily at ECoG electrodes 3, 4, and 5 and exhibited the shape characteristics of a prototypical SD as shown in Figure 2A.

Of these 140 ECoG-coded SDs and ISDs, 39 SDs could be visualized in both the Prototype device channel plots and the movies, but just 26 of these were deemed readable in the Prototype device recordings. Notably some of these 26 epochs exhibited minimal artifacts during their DC-shifts but were still deemed readable. Gaussian plus baseline curve fitting with 4-min epoch widths was performed for 24 of these 26 DC-shifts, with 2 epochs fitted with 12 min widths. The parameter fitting errors of all of the 26 Prototype device and 26 ECoG peak signal epochs were under 11%, although in one of these 26 epochs some of the 8-sec bin values were removed due to obvious artifacts.

### 3.3 DC-shift comparison and characterization

The CoD of corresponding 8-second segments from all 26 of the ECoG and CerebroPatch™ Proof-of-Concept Prototype DC-shifts was calculated to enable their comparison. The histogram of these CoDs is shown in Figure 3A. Two of these 26 epochs had CoDs less than 0.80 thus excluding them from further analysis based on a lack of similarity. The CoD of one of these was 0.797, because noisy portions of the time courses were excluded from the fitting procedure. This process left 24 DC-shifts that were deemed structurally similar. The median [IQR] CoD of the 24 scaled Prototype device and ECoG DC-shifts that met the 0.80 limit of similarity was 0.98 [0.02], n = 24.

**Figure 3.**
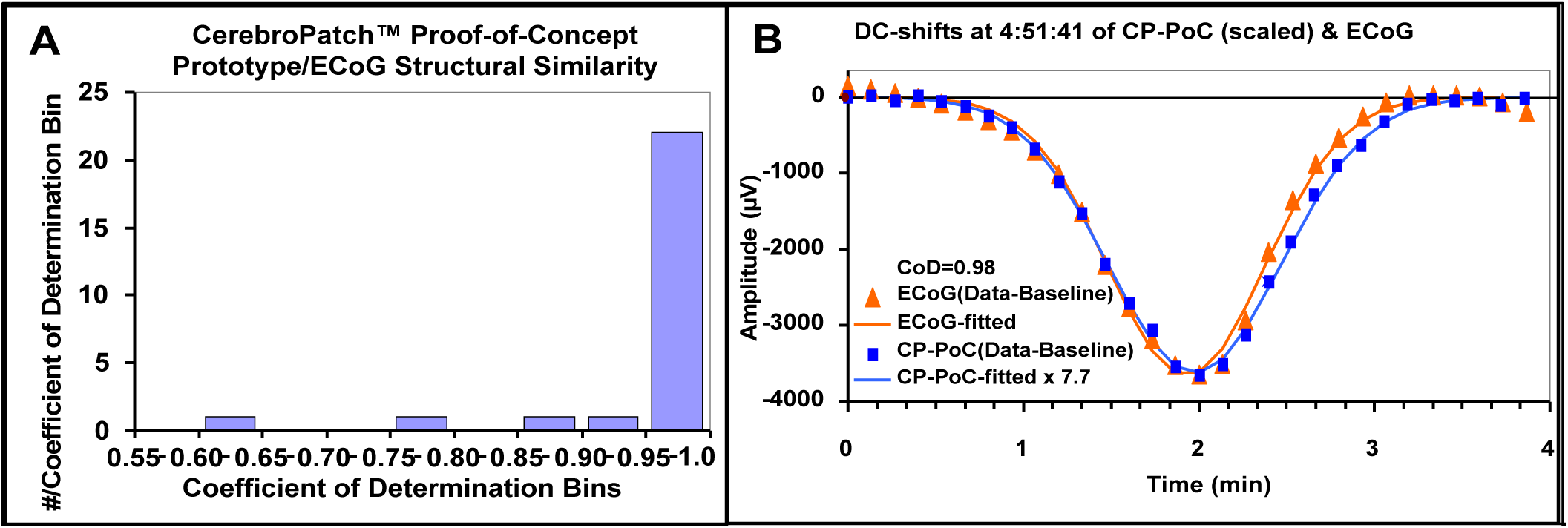
Panel A shows the frequency distribution of the 26 Coefficients-of-Determination (CoD) between the 8-sec data segments of ECoG strip and CerebroPatch™ Proof-of-Concept Prototype device curves showing the two excluded CoDs of 0.63 and 0.79 whose curves were deemed not similar. The remaining twenty-four DC-shift comparisons with CoDs >0.80 suggest strongly that the Prototype device detected the confirmed ECoG-coded SDs. In Panel B the data from 3^rd^ SD shown in Figure 2A and 2B is shown. The 8-sec data block averages of the ECoG (red triangles) and CerebroPatch™ Proof-of-Concept (CP-PoC) Prototype (blue squares, scaled by the 7.71 ratio of the peak signal amplitudes) signals of the largest DC-shifts are fitted to a 6 parameter baseline plus Gaussian model that show their structural similarity evidenced by a CoD of 0.98.

The CerebroPatch™ Proof-of-Concept Prototype-visible DC-shifts were characterized by the Gaussian fit parameters (see Table 2) including the peak DC-voltages of -457 ± 69 µV, n = 24, in comparison to the ECoG voltage of -3771 ± 171 µV, n = 24, with a voltage ratio of 0.121 ± 0.016, n = 24. The duration expressed as the FWHM of the fitted curves of the ECoG and Prototype device DC-shifts was 63.2 ± 3.65 sec, n = 24, and 70.9 ± 5.92 sec, n = 24, (p<0.001, paired t-test), respectively, with the Prototype device duration being longer presumably because of the spread of the electric field as it travelled through the layers from the brain to the scalp. The time of the Prototype device peak was delayed compared to the ECoG peak by a median of 2.89 [2.32] sec, n = 23 sec (with one preceding removed). This delay was much less than the sampling time of 8 seconds and corresponds to 4.3% of the average duration.

### 3.4 SD cluster and structural comparison

The raw DC-potential data from the ECoG strip and CerebroPatch™ Proof-of-Concept Prototype from a cluster of 5 DC-shifts are presented in Figure 2A and 2B, respectively, after being down-sampled into 8-sec data blocks. This cluster of five DC-shifts was part of a cluster of 25 DC-shifts that occurred over a period of 8.25 hr with a mean interval of 22.4 ± 0.4 min, n = 24. Their DC-shifts appeared in a similar configuration of Prototype device electrodes. Figure 3B shows the structural similarity of the processed data derived from the 8-second binned raw ECoG and Prototype device data of the 3^rd^ of the 5 DC-shifts presented in Figures 2A and 2B. This structural similarity is demonstrated by a CoD of 0.98.

### 3.5 Spatial extent

Although the time-courses of the ECoG and CerebroPatch™ Proof-of-Concept Prototype DC-shifts match at the electrode level, the brain-surface DC-potential of the SD spatially spreads as it travels through the various tissues to the scalp [7]. The wide spatial spread of the Prototype device DC-shifts on the scalp was characterized by an area of 18.7 ± 2.76 cm^2^, n = 24, that encompassed the region within the 50% isocontour and a diameter of a circle with the equivalent area of 4.87 ± 0.365 cm, n = 24. The 4-5 cm extent of the scalp DC-shift at the peak at 4:51 min:sec is shown in Prototype device image in Figure 2C.

### 3.6 CerebroPatch™ Proof-of-Concept Prototype video

For this same epoch of 5 DC-shifts shown in Figures 2A and 2B, Supplemental Video S1 shows the ECoG and Prototype device channel plots, their similar time courses, and a heat-map movie of all 5 of the DC-shifts with their rapid depression and recovery with FWHMs of ∼65 sec and peak voltages that approach -410 µV. The extent of this DC-shift for the 3^rd^ SD includes Prototype device electrodes A4, A5, B4, B5, B6, C5, C6, C7, D4, and D5 and covers an area of 20 cm^2^ with its associated diameter of 5 cm at its maximum deviation time of 4:51 min:sec.

## 4. Discussion

### 4.1 Noninvasive SD detection is possible using scalp DC-potentials

The main findings of this study are: 1) artifact issues limited the analysis to 10% (0.85/8.31 days) of the recording time in one patient and did not allow data analysis in the other 2 Cohort 2 patients with ECoG-identifiable SDs; 2) Usability was severely limited by the necessity of the re-gelling procedure and the inadequacy of the scalp adhesion system; 3) the DC-shifts of brain-surface SDs can be detected from the scalp; 4) the processed and scaled scalp DC-shifts from the CerebroPatch™ Proof-of-Concept Prototype and those from the gold-standard ECoG SDs are almost identical as indicated by their matching curve parameters and CoDs greater that 0.80; 5) the extent of the scalp DC-potential is consistent with its spread during its passage from the brain surface to the scalp as estimated using numerical simulation; and 6) the scalp/brain voltage ratio is higher than previous estimates. These findings are tempered as they occurred in one subject with one specific and unusual SD pattern on ECoG consisting of large nearly isoelectric DC-shifts that spread to adjacent channels as both DC-shifts and EEG suppression.

The structural similarity of the invasively recorded DC-shifts identified as SDs from the gold-standard ECoG method [33, 34] and the noninvasively recorded DC-shifts from the CerebroPatch™ Proof-of-Concept Prototype was assessed by a processing procedure that allowed for their quantitative comparison. The similarity of 24 of 26 DC-shifts with a CoD greater than 0.80 is documented by a median [IQR] CoD of 0.98 [0.02] as presented in Table 2 with an example shown in Figure 3B. This numerical comparison of the invasively and noninvasively detected DC-shifts and the visualization and spatial characterization as enabled by our closely-spaced 1-cm electrode array distinguishes our results from previous scalp DC-shift recordings [6, 39]. Their more widely spaced scalp electrode placements would have made the reconstruction and visualization of the scalp electric field and the optimal alignment of the scalp and brain surface electrodes for the quantitative comparison of their DC-shifts more difficult.

This quantitative evidence of similarity gives support to the suggestion that this proof-of-concept validation data of noninvasive SD detection can lead to the implementation of clinically useful SD detection. If the artifact and usability issues of the proof-of-concept Prototype device issues can be corrected in a future design which would then be validated against ECoG, artifact-free and user-friendly noninvasive SD detection using DC-EEG could be used for evaluating brain-saving SD-based therapies to improve outcomes in sABI patients by mitigating the effects of secondary brain injury.

### 4.2 Scalp/brain DC-shift voltage ratio

The results of our numerical simulation of SDs [7] were helpful in associating the scalp DC-potentials with ECoG coded SDs. However our scalp/brain voltage ratio of 0.121 is larger than our previous values of 0.0735 from concussive-SD numerical simulation [7] and 0.0316 from Drenckhahn et al.’s [6] SAH patient data presented by Hund et al. [7]. The differences between these ratios can be attributed to several factors, including SD morphology differences (as presented previously [7]), skull thickness variations, brain/scalp electrode misalignment, and differing proximity to possible bone defects.

The scalp/brain voltage ratio of an SD is highly dependent on skull thickness. Our simulation results showed that 44% of the voltage drop between the brain surface and the scalp occurs due the resistivity of the skull [7], so variations in skull thickness have a major effect of the voltage ratio. From the simulation data of Hund et al. [7], the ratio would increase from 0.0735 to 0.109 (by 49%) if the estimated skull thickness of 5.7 mm was reduced by 0.5 mm (by 9%), emphasizing how important skull thickness and its resistivity is to the scalp/brain ratio. Skull thickness is highly variable both within [55] and between individuals with coefficients of variation from 20 to 32% [56, 57].

The Drenckhahn et al. [6] ratio of 0.0316 reported in Hund et al. [7] was from a comparison of various locations of brain surface and scalp electrode positions that were not aligned to provide positions that sampled equivalent regions of the SD’s voltage field at the brain surface and scalp. In addition, the skull thickness differences from the various scalp electrode positions could potentially result in more resistive lowering of the scalp voltage than for our data, and therefore a lower voltage ratio, whereas our ratio of 0.121 was calculated from the maximum voltages from each electrode location, be it brain surface or scalp.

### 4.3 Noninvasive SD identification criteria

The exploratory SD-identification method used here involves the quantitative comparison of fitted DC-shift curve parameters from channels with maximum voltages combined with the representation of the spatial distribution of the scalp DC-potential. We justify our focus on DC-EEG for noninvasive SD detection because of its importance in identifying the primary attribute of SD and because DC-based SD detection is important during ISDs when the detection of the suppression of high-frequency signal is not useful. The importance of DC-EEG for SD identification is in contrast to its lack of acceptance for diagnostic clinical EEG. Our approach of using DC-EEG to explore noninvasive SD detection of the primary DC-shift of an SD differs from several efforts to explore noninvasive SD detection using clinical frequency EEG with 2.5 cm or greater electrode spacing that have focused on providing an algorithm to assess the width and duration of SD’s secondary EEG suppression as SD identification criteria [40, 41, 58].

### 4.4 Observational limitations

The limitations of this proof-of-concept study of noninvasive SD detection include not observing DC-shift propagation or EEG suppression in the DC-shift scalp and clinical EEG frequency recordings, respectively. This lack of two of the three invasive ECoG-based criteria of SD coding and identification set forth by COSBID [33, 34] is supported for noninvasive SD identification by Hartings et al. [43] who stated that,

> “[N]o criteria for definitive diagnosis of SDs by scalp EEG has yet been proposed. [Although the] criteria for ECoG recordings from the brain surface using subdural electrodes … have been firmly established [33] [it] is unlikely that these criteria could also be applied in a straightforward manner for SD identification in scalp EEG.”

### 4.5 Lack of propagation

The Prototype device movies created from Gaussian radial-basis function interpolation of the 2D array of DC-potentials [53, 54] were capable of visualizing potential propagation, but did not provide evidence of propagation. The Prototype device image movies do show DC-shifts that are synchronous with the ECoG DC-shifts that were coded as SDs by an experienced reader. These DC-shifts appear in one contiguous region and then dissipate without apparent propagation in scalp recordings, but clearly demonstrated propagation on the ECoG recordings. Perhaps propagation could not be observed in the scalp recordings because the SD moved out of the region covered by the Prototype device’s electrode array.

There is a possibility that the lack of scalp DC-shift propagation did actually reflect cortical non-spreading depolarizations for some of our data. This conjecture is supported by observations of cortical non-spreading depressions [31] and depolarizations [20, 45]. In the first human observation of spreading depression with the ECoG strip methodology [31], 20% of the 48 spreading depressions were estimated to be “stationary” based on subtracting the estimated number that would be deemed as to have travelled perpendicular to the ECoG strip. Dohmen et al. [20] specifically mentions not reporting 65 slow potential changes associated with SDs that “did not show clear spread of depolarizations” to abide by the criteria that “specify spread of depolarization as a prerequisite” for scoring an SD. Bastany et al. [45] also observed “stationary” cortical depolarizations. We surmise non-propagating SDs are underreported by investigators because they are presumed to be traveling perpendicular to the ECoG strip, are classed inappropriately as artifacts, or are dismissed as missing one of the COSBID-required essential identifiers of SDs.

### 4.6 Lack of EEG suppression

Although several attempts were made to replicate the EEG suppression observed in the ECoG recordings in the CerebroPatch™ Proof-of-Concept Prototype channel plots and movies, we were not able to achieve this analysis, thus exposing a limitation that the scalp DC-shifts detected by the Prototype device cannot be characterized as ISDs. Replicating the bipolar analysis used to improve the observation of EEG suppression in ECoG recordings from the multiple signals in a closely-spaced 2D electrode array was attempted by subtracting high- and low-isocontour-selected Prototype device electrode signals but did not produce meaningful results.

### 4.7 Number of observations

For this validation study SDs are the object/event to be validated, not the patient, so our “n” is not “one” as the number of subjects used for data, but 26 as the number of SDs. Even though these 26 SDs were from the same subject, we justify our use of the SD as the independent observational unit because SDs that emanate from sABIs that are classed as “transient, short lasting” [59] all have the same time-course attributes based on the unity of their bioelectrical and biophysical attributes. This rational of the number of observations for this validation study is distinct from the situation for most clinical studies where the object being evaluated/analyzed is the patient.

### 4.8 Device-design based limitations: Impact of artifacts and usability on SD identification

The CerebroPatch™ Proof-of-Concept Prototype’s ability to gather artifact-free data was severely limited. Many of the 140 ECoG-coded SDs in this data set were inaccessible for testing scalp SD detection due to inherent limitations in the design of the Prototype device. The excessive artifact burden in our data could be viewed as a severely limiting negative result. Fully acknowledging this negative data to avoid unnecessary future studies as a scientific principle is slightly analogous to the entrepreneurial dictum “fail faster to succeed sooner”. This failure of the prototype device suggests specific design features for a new future device. A different device design could well eliminate or minimize the Prototype device-based limitations of artifact burden and lack of usability. From this point-of-view, reporting the artifact burden removes, rather than enhances, the preliminary aspects of our study. This artifact burden suggests that additional of proof-of-concept studies would be necessary to validate a re-designed DC-EEG system that would improve usability and reduce artifacts. Additional observations to validate the new design might enable observation of DC-shift propagation and EEG suppression at the scalp and confirm the DC-shift-based proof-of-concept observation presented here that the DC signal from closely-spaced scalp-placed electrodes could provide noninvasive SD detection.

### 4.9 Effect of ECoG strip on SD voltage field propagation

Our unpublished numerical simulation of an SD passing under an ECoG strip showed that the surface signal is increased by 19% when passing under the platinum electrodes in the ECoG strip or diminished by 8% when it passes under their silastic encasement. These simulation results suggest that SDs can be detected even if they pass under the ECoG strip and that the increase in signal when the SD passes under the platinum electrode portion of the ECoG strip was not large enough to suggest that our SD detection was dependent on this increase. Therefore, we are confident SD detection by the CerebroPatch™ Proof-of-Concept Prototype was not dependent on, or inhibited by, the presence of conductive properties of the ECoG strip, nor obscured by its resistive elements.

### 4.10 Future device design recommendations

As this study presented negative results concerning artifact burden and limited usability, we feel it imperative that these negative results be used to provide positive recommendations for future design criteria and prototyping procedures. This is important to further the goal towards clinically useful noninvasive SD detection with the object of minimizing the detrimental effects of secondary brain injury by developing SD-targeted therapies and then monitoring their effectiveness.

We recommend that the protocol structure of this proof-of-concept validation study be retained for the next development stage. This protocol structure with 3 Cohorts would have performed adequately if the CerebroPatch™ Proof-of-Concept Prototype had been properly designed. However, to make sure a future device is properly designed, we envisage an iterative design [60, 61] prototyping design procedure that allows for multiple prototypes in stages of increasing complexity. This iterative design prototyping system permits the redesign of ineffectual and underperforming design elements before the next prototype stage is attempted, thus avoiding the “build it, then send it” system that was used to design the Prototype device.

We suggest that effective noninvasive SD detection in the neuro-ICU environment needs an easily applied electrode array design. An important feature of our CerebroPatch™ Proof-of-Concept Prototype device, that the electrode array was positioned and held in place as one item, should be carried over to a new device as this eliminates the sequential placement of many single electrodes, a time-consuming, labor-intensive, and user-unfriendly practice. This array should include provisions for robust scalp adhesion that provides a stable skin-electrode interface to minimize artifacts but that maintains the minimal skin-irritation feature of the Prototype device, wireless signal transmission to avoid cable-generated artifacts and to ease removal and replacement for imaging and other procedures, provisions for hair penetration to increase recruitment possibilities and remove the limitation that SD-generating lesions overlap the forehead.

## 5. Conclusions

Because of the limitations of this proof-of-concept study, we are able to provide future device design recommendations. Despite these limitations, we showed that the time-matched CerebroPatch™ Proof-of-Concept Prototype scalp DC-shifts originate from SDs validated using the COSBID identification criteria [33, 34]. In addition, the spatial characteristics of the CerebroPatch™-detected DC-shifts are consistent with the spread of the brain-surface DC-potential from a SD. These results suggest that the Prototype device DC-shifts are from SDs and that noninvasive SD detection is possible using scalp DC-potential signals. Although the Prototype device SD identification procedures depended on SD identification using the ECoG strip gold-standard, they did enable the proof-of-concept validation that the Prototype device was capable of identifying SDs.

## Data Availability

All data produced in the present study are available upon reasonable request to the authors based on the submission and suitability of a formal project outline.

## Acknowledgements

We are grateful for the technical assistance provided by Nicholas Urioste, Amal Alchbli, and Mohammad Abbas and to Mihika Gangolli for the suggestion to use the Coefficient-of-Determination to compare DC-shifts. We thank Martin Fabricius, Thomas Ferguson, Rebekah Kummer, and Joel Greenberg for reading and commenting on the manuscript. We acknowledge support from the Ansys StartUp Program and the Duquesne University Small Business Development Center.

## 6. Author contributions

ELS contributed to the design, construction, and maintenance of the CerebroPatch™ Proof-of-Concept Prototype system. BRB developed the analysis code with input from SCJ and SJH and wrote the first draft. BRB contributed additional analysis. KAE conceived of and performed the statistical analysis.. SCJ, BRB, CWS, and APC edited the manuscript. SCJ conceived of the project, provided supervision, and revised the manuscript,

## 9. Compliance with Ethical Standards

All ethical standards have been met. See section “Human study issues”.

## 10. Data and Code Availability

Available upon request based on the submission and suitability of a formal project outline.

## 11. Intellectual Property Disclosures

Parts of this work are disclosed in: U.S. Patent No. 10,028,694 published and publically available on May 26, 2016 and issued July 24, 2018; U.S. Patent No. US11234628B2 issued February 1, 2022; and European Patent No. 3223693 issued February 28, 2024.

## 12. Supplementary material

Supplementary material is available at the journal’s website.

## 15. Supplemental Material

### 15.1 Supplemental Video 1

Supplemental Video 1: This video shows the reconstructed heat-map movie from the CerebroPatch™ Proof-of-Concept Prototype channel electrodes voltages for a patient with aSAH (upper left panel) with the voltage calibration pseudo-color scale from 250 to -410 µV (positive-negative reversed) just below. A frame of this video at 4:51 was depicted in Figure 2C as a still image. The extent of the DC-shift scalp voltages is depicted in the heat-map movie as “yellow” ∼4 cm regions of approximately -410 µV in the upper right corner. The channel plots of the ECoG electrodes are shown in the lower left panel and those of the Prototype device in the right panel with the time scale below. The electrodes in Prototype device image are represented as small black circles labeled in five rows with 5 electrodes in rows A and E, 6 in rows B and D, and 7 in row C with a filled-circle depicting electrode A1. The vertical red line is a time-marker that moves across the channel plots as shown on the time-ticker just below the movie frame created from the Prototype device data.

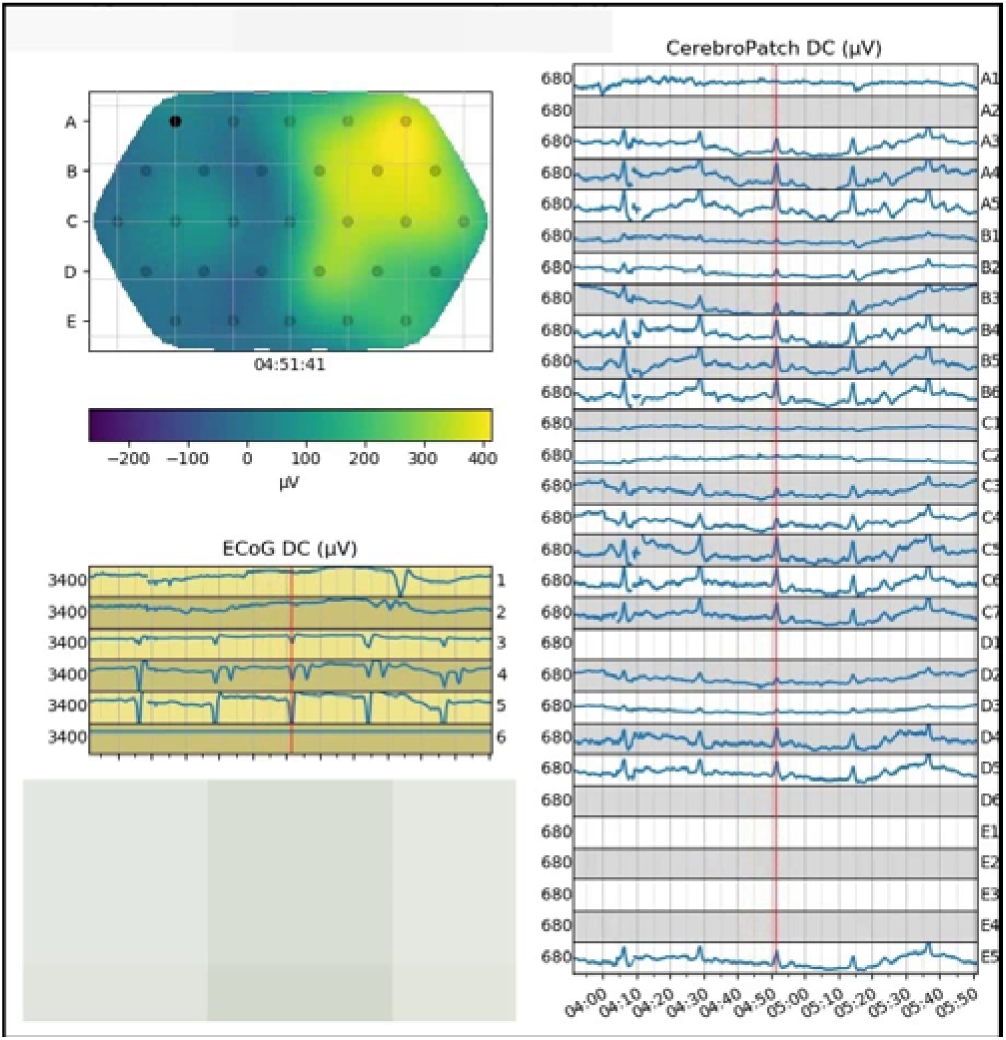

